# Seizures may worsen outcomes of neonatal hypoxic ischemic encephalopathy: a longitudinal serum biomarkers study

**DOI:** 10.1101/2024.05.20.24307562

**Authors:** Khyzer B Aziz, Jordan Kuiper, Alison Kilborn, Hrishikesh Kambli, Srishti Jayakumar, Gwendolyn J Gerner, Aylin Tekes, Charlamaine Parkinson, Ernest M. Graham, Carl E. Stafstrom, Christopher Campbell, Catherine Demos, Martin Stengelin, George Sigal, Jacob Wohlstadter, Allen D. Everett, Frances J. Northington, Raul Chavez-Valdez

## Abstract

**Introduction:** Screening and treatment of seizures (Sz) in neonates suffering from hypoxic-ischemic encephalopathy (HIE) is routine. Understanding if Sz worsen brain injury and outcomes would optimize treatment decisions, recognizing risks and benefits. We hypothesized that serum central nervous system (CNS)-specific biomarkers would discriminate neonates with Sz and relate to outcomes.

**Methods:** This retrospective cohort study was conducted between April 28, 2009 and November 15, 2019, including neonates diagnosed with seizures (Sz) and/or HIE treated with therapeutic hypothermia (TH), who had sufficient remnant serum for biomarker analysis. Neonates were grouped in i) only Sz without HIE (Sz-no HIE), ii) HIE with Sz (Sz-HIE) and iii) HIE without Sz (no Sz-HIE). Levels of glial fibrillary acidic protein (GFAP, astrocytic reactivity), Tau (neuronal injury) and neurofilament light chain (NF-L, axonal degeneration) were studied at admission, <72h and 72-144h of life against time to full oral feeds and brain injury in MRI.

**Results:** After exclusions 145 neonates were included (61% male; 33% Black). Admission GFAP levels were higher in Sz-HIE than in no Sz-HIE neonates. During the first 72h of life, levels of all 3 biomarkers were similar between Sz groups (Sz-no HIE & Sz-HIE), but higher than in no Sz-HIE. After 72h, NF-L and Tau remained higher in both Sz groups vs. no Sz-HIE. Stroke diagnosed in 31% (Sz-no HIE), 8% (Sz-HIE) and 11% (no Sz-HIE) of neonates had no effect in biomarkers levels. In adjusted regression models higher Tau and NF-L percentiles related to longer time to reach full oral feeding and higher odds of significant brain injury in MRI in both Sz groups.

**Conclusions:** In this study, Tau and NF-L levels are higher in those neonates developing Sz, irrespective of their HIE diagnosis. These results provide additional support for the notion that Sz may worsen brain injury and outcomes in neonates with HIE even with TH.

## INTRODUCTION

Hypoxic-ischemic (HI) encephalopathy (HIE) is the most frequent cause of neonatal encephalopathy (NE).^1^ Although, longitudinal neurological exams and electroencephalographic (EEG) evaluations are among the most valuable prognostic tools available, predicting the outcomes of HIE survivors in the era of therapeutic hypothermia (TH) still remains challenging, as many interventions (i.e., sedation, anti-epileptic drugs) may interfere with these assessments. Thus, there is a need for reliable biomarkers to better stratify patients to future adjuvant therapies and predict outcomes.

The immature brain has a higher susceptibility to Sz compared to adults, due to factors modulating the brain’s excitation / inhibition balance (i.e., depolarizing GABA,^2–4^ higher expression of NMDA-type glutamate receptors,^5^ earlier expression of excitatory ion channels^6^). Although early electrical seizures (Sz) are used as a marker of moderate to severe HIE, the relationship between Sz and worsen brain injury remains poorly understood. For instance, Sz burden has been linked to worse neurodevelopmental deficits in HIE;^7–9^ however, the question remains if Sz by worsening brain injury result in poorer outcomes. Even when Sz during HIE are often the result of acute inflammation and thus, prolonged treatment is not usually necessary, knowing if Sz may worsen brain injury is important to optimize treatment decisions, recognizing the risks and benefits of anti-seizure drugs.^10^

Here, we aimed to measure GFAP (astrocytic activation), Tau (neural cytoskeletal damage), and NF-L (axonal degeneration) in the serum of neonates with either HIE and/or Sz hypothesizing that serum central nervous system (CNS)-specific biomarkers would discriminate neonates with Sz and relate to short-term outcomes.

## METHODS

### Design and setting

Single-center, retrospective cohort study conducted at a tertiary academic level IV NICU.

### Participants & Study Size

This study had exempt status from the Johns Hopkins University (JHU) Institutional Review Board (IRB), until 2017, when informed consent became required. The study (IRB00026068 and IRB00333123) complied with the Health Insurance Portability & Accountability Act of 1996 (HIPAA). STROBE criteria was followed for report of results.

#### Inclusion criteria

Neonates diagnosed with either HIE treated with TH and/or electrical Sz were included to compared them in 3 groups: i) Sz without HIE (Sz-no HIE), ii) Sz in the setting of HIE treated with TH (Sz-HIE) and iii) HIE without electrical Sz (no Sz-HIE).

##### HIE diagnosis and TH initiation

Our institutional guidelines to diagnose HIE requiring TH were adapted from the National Institute of Child Health and Development (NICHD) - Neonatal Research Network (NRN) criteria.^11^ TH was started within the first 6h of life in neonates of ≥ 1800 g of birth weight (BW) and ≥ 35 weeks gestational age (GA) if: a) cord or 1^st^ hour of life pH ≤ 7.0 and/or base excess (BE) ≤ −16 or b) cord or 1^st^ hour pH > 7.0 but ≤ 7.15 and/or BE was between −10 to −15 plus evidence of a sentinel event and either need for assisted ventilation or Apgar < 5 at 10 min of life. Although persistent or worsening moderate to severe encephalopathy (modified Sarnat scores of 2 or 3 ^12^) during the first 6h of life is a necessary criterion to initiate TH, around ∼20% of HIE neonates included in this analysis who received TH had only signs of mild encephalopathy (Sarnat score of 1). These neonates were deemed by the treating physician to be at greater risk to develop worsening neurological exam after the first 6h of life based on clinical history and biochemistry.

##### Sz diagnosis

Until 2014, routine (1h) EEG along with continuous amplitude-integrated EEG (aEEG) were used in HIE infants undergoing TH instead of our current 72h full-montage video-EEG. However, all neonates evaluated for suspected seizures received a full-montage video-EEG for 6-48h if HIE was not suspected. Thus, for our study we classified Sz as those with EEG confirmation or sufficient concern by the pediatric neurologist to maintain anti-seizure therapy in the setting of an abnormal EEG background and concerning aEEG. Of a total of 290 screened neonates diagnosed with HIE and/or Sz, 44 of them were diagnosed with Sz but not HIE (Sz-no HIE) and 246 were diagnosed with HIE (with or without Sz). Of these, all 44 Sz-no HIE and 130 HIE neonates had enough remnant serum samples to run biomarker analysis (**Flowchart**).

#### Exclusion criteria

Of 174 babies who met inclusion criteria with biomarker data available. 29 were excluded (16.6%) due to: i) < 72h of TH for HIE (7, 4.0%), ii) genetic/ congenital disorder (6, 3.4%), iii) Extracorporeal membrane oxygenation (ECMO; 13, 7.5%), iv) < 35 weeks GA (5, 2.9%) and v) <1800 g BW (1, 0.6%). Two excluded babies with HIE who underwent ECMO also had a genetic disorder or were cooled for <72h. One neonate diagnosed with HIE was excluded for receiving TH for <72h and being <35-week GA. Following exclusions, 95.5% (42 out of 44) neonates in Sz-no HIE group and 79.2% (103 out of 130) neonates with HIE (36 Sz-HIE and 67 no Sz-HIE) were included. Of a total of 246 neonates with HIE/TH, 190 remained after exclusions (77.2%) and 103 of them had serial biomarker data (54.2%). The percent of outborn neonates and the base deficit were the only differences identified between those HIE/TH neonates included or excluded from the analysis (**Supplemental Table S1b**). Although, 9 out of 42 neonates (21.4%) included in the Sz-no HIE group had evidence of mild encephalopathy (Sarnat score of 1) at admission, none of them were diagnosed with hypoxic-ischemic injury (HIE) and NE was attributed to other causes (i.e., stroke), as none of the HIE diagnostic criteria (i.e., clinical history, biochemical data) were identified.

### Variables

#### Independent variables

Clinical data compared between groups are included in **Table 1**. Race was assigned based on self-identified maternal race. Sex at inspection, GA, and Apgar scores were determined by the NICU team. Serial neurological exams were performed by pediatric neurology and treating teams at least twice a day for all neonates in the study.^13^ However, in contemporary cohorts many factors (i.e., sedation during TH, hemodynamic instability, anti-seizure medications) may decrease the expected positive predictive value of these exams during the sub-acute phase of illness, therefore for this study only the highest modified Sarnat score during the first 6h upon NICU admission, entry criteria for NICHD-NRN TH RCT,^11,14^ was documented for analysis. Other clinical definitions included: i) multi-organ failure: failure of at least 2 organ systems in addition to the neurological system; ii) renal failure: oliguria (< 0.5 ml/k/h of urine output) beyond 24h of life with increasing creatinine levels; iii) hepatic failure: worsening transaminitis with albumin <3 g/ dL and/or prolonged prothrombin time (>16 seconds); iv) cardiovascular failure: cardiac dysfunction with decrease cardiac output by echocardiogram and/or systemic hypotension and/or pulmonary hypertension; v) pulmonary/ventilatory failure: need for supplemental oxygen and/or ventilatory support via invasive or non-invasive methods and vi) hematological failure: decrease in any of the 3 main blood cell lines and/or disseminated intravascular coagulopathy (DIC). The worst (lowest) pH and BE measures (cord or 1^st^ BG) were used for analysis, as they better represent clinical practice. Neonates undergoing TH had a head ultrasound (HUS) prior to initiation and immediately after completion of TH. Diagnosis of stroke was made by MRI and categorized as middle cerebral artery (MCA) or any. Serum remnants samples were collected daily from day of life (DOL) 0 to 6 and stored at −80°C. Each aliquot was exposed to only 1-2 thaws/freeze cycles prior to assay. After diluting samples 5-fold, an ultrasensitive multiplex enzyme-linked immunosorbent assay (ELISA) was used to measure GFAP, Tau, and NF-L in 96-well plate (Meso Scale Discovery [MSD], Rockville, MD).^15^ For biomarker values below the lower level of detection (< LLOD), we used LOD/√2 as recommended,^16^ while for those above we use 10x the upper LOD (> ULOD).

**Table 1.**
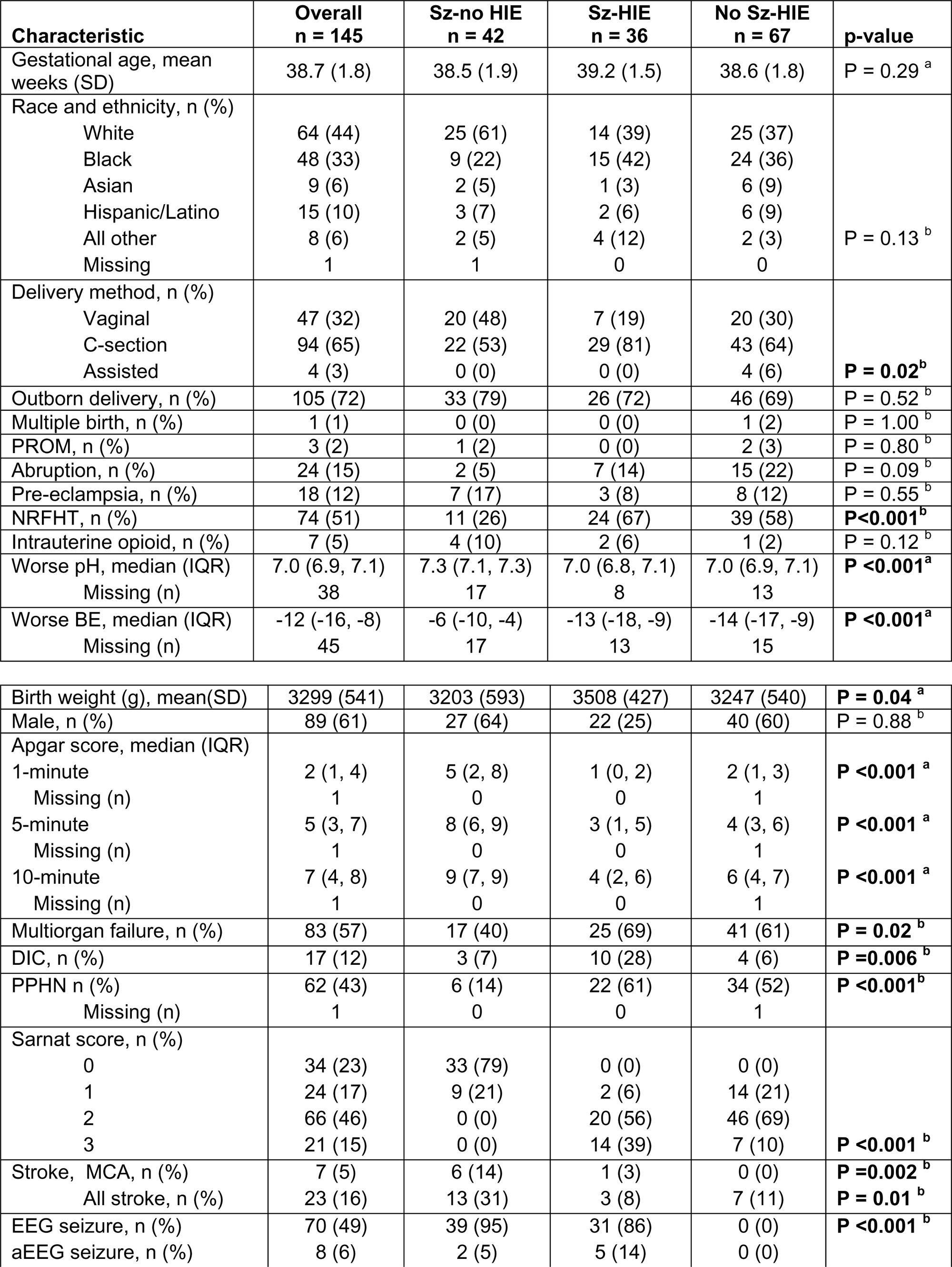
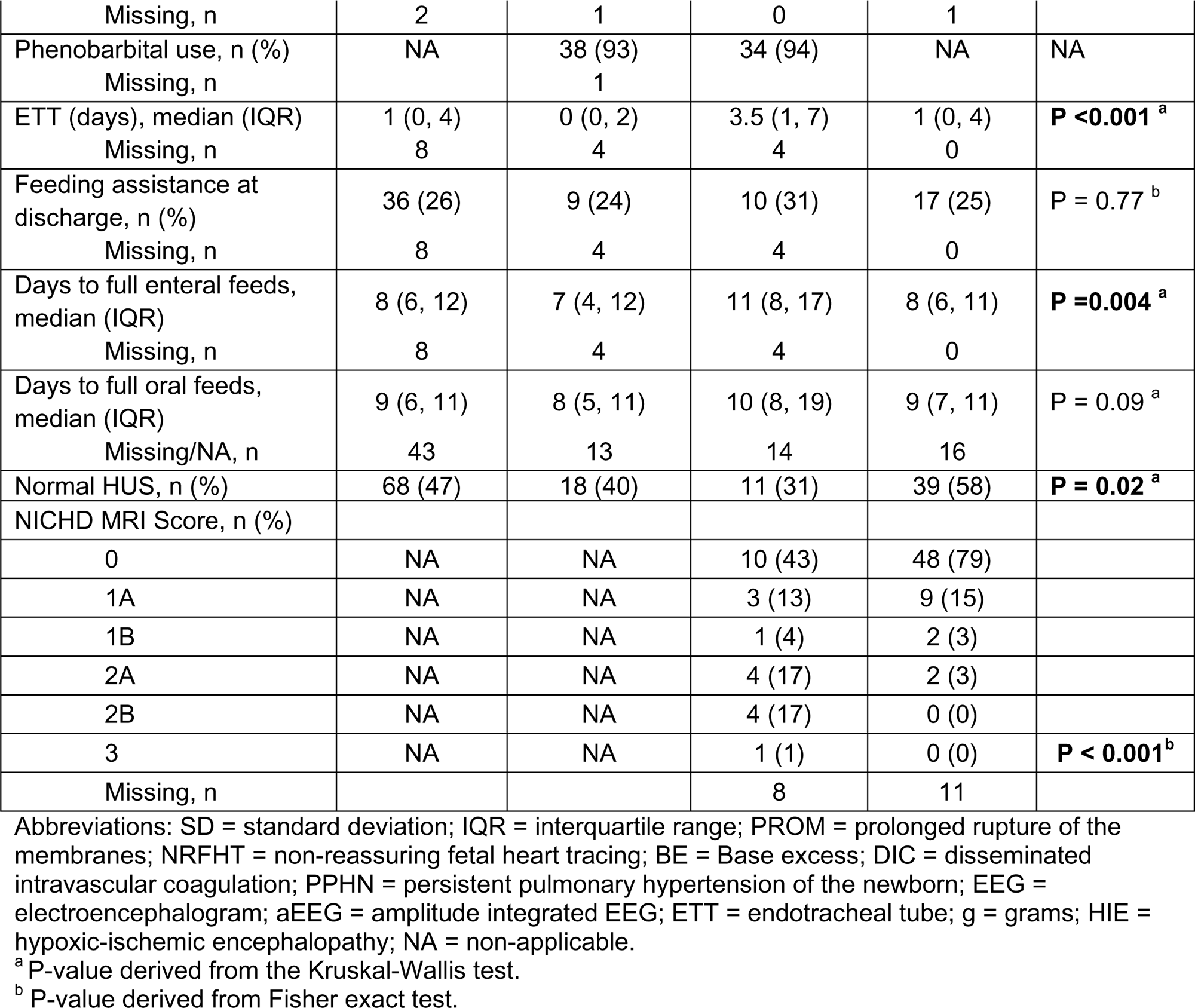
Descriptive statistics of maternal and patient characteristics, by group.

#### Outcome variables

Neurological recovery and injury variables included impaired oral feeding (for all 3 groups)^17^ and significant brain injury by NICHD-NRN MRI scale (for HIE groups)^18^. Feeding data collected included: i) days to full enteral feeds (day of parenteral nutrition discontinuation, by protocol at ≥120 ml/k/d), ii) days to full oral feeds (day of gavage tube discontinuation) and iii) need for feeding assistance at discharge (i.e., gastrostomy tube). Brain MRI was performed using 1.5 or 3T magnets as soon as feasible upon completion of rewarming (IQR, 5-9 DOL). The NICHD NRN score system classifies patterns of brain injury using T1 and T2 weighted MRI.^18^ Levels 0 (normal) and 1A (minimal cerebral lesions without involvement of basal ganglia, thalamus or internal capsule or watershed infarction) were classified as “normal-minimal injury” and anything beyond (1B, 2A, 2B or 3) were classified as “significant injury” for modeling. This dichotomization was used based on the known associations between moderate-severe neurodevelopmental impairment in <5% of neonates with levels 0 and 1A and between 25 to 100% in those above 1A.

### Data sources

Clinical and biochemical data (**Table 1**) were obtained from electronic records. Assignment of the highest modified Sarnat score during the first 6h upon NICU admission was determined by RCV, CP and FJN. Biomarker analysis was performed at ADE laboratory. EEG evaluation and Sz assignment was performed by pediatric neurologist (CS). HUS assessment and MRI NICHD NRN scoring was performed by neonatal neuroradiologist (AT).

### Statistical Analysis

Descriptive statistics were calculated for patient-level variables and biomarkers in the overall study population and by each of the 3 clinical groups. Several continuous variables were not normally distributed; thus, statistics were summarized as median and interquartile range (IQR). To test for differences in categorical, patient-level variables across the 3 clinical groups, Fisher’s exact tests were applied. For continuous patient-level variables, non-parametric statistical methods were used including Mann-Whitney U (MWU) or Kruskal-Wallis H-test (KW), with Conover-Iman tests with Bonferroni correction for pairwise comparisons across the 3 groups. The same methods were used to test for differences in biomarker concentrations across the 3 groups, separately by timepoint (i.e., cross-sectional tests of differences across groups). To describe and characterize differences in median biomarker concentrations over adjacent timepoints, separately within each of the 3 groups, quantile regression with cluster-robust standard errors was used (i.e., longitudinal tests of differences within groups). To evaluate the covariate-adjusted associations of each biomarker with feeding outcomes, a modified Poisson regression censored at the 95^th^ percentile (21 days) was used. Censoring was used to avoid associations driven by a small number of outliers. For models of MRI-based brain injury scores, two different approaches were used. First, we used an ordinal logistic regression model to estimate the (cumulative) odds ratios (and 95% CI) associated with a doubling in biomarker levels. These models provide a single odds ratio for being in a higher brain injury severity level, assuming proportional odds across categories. Second, penalized logistic regression was used with dichotomized outcomes with cutoffs at >0 (normal & abnormal) or >1A (normal-mild injury & significant injury) levels per doubling in brain injury biomarker concentration. All models were adjusted BW, delivery mode, and multi-organ failure. For models evaluating associations of biomarkers, levels from 24 to <72 hours and 72 to 144 hours were simultaneously included to allow for co-exposure adjustment. Lastly, machine learning methods were apply to calculate Shapley Additive Explanations (SHAP) values to determine which features were most important in regression modeling to prevent over-fitting. A p-value of ≤ 0.05 was considered significant. GraphPad Prism (Version 8.0.0(131) 2018; GraphPad Software, San Diego, CA), Stata (Version 15, StataCorp LLC, College Station, TX), and Python (Version 3.7) software were used for analysis.

## RESULTS

### Descriptive data

#### Patient characteristics

After exclusions, 145 neonates were included in analysis with all characteristics shown in **Table 1**. Neonates in the Sz-HIE group were heavier, more often delivered via C-section and more likely to develop multiorgan failure (including DIC and PPHN) compared to those in the Sz-no HIE and no Sz-HIE groups (**Tables 1** & **S1a**). Neonates in both HIE groups had more often history of NRFHT and lower Apgar scores, cord pH and base deficit than those in the Sz-no HIE group (**Table 1**). Strokes occurred in 31% in the Sz-no HIE group, while only in 9.7% in the HIE groups (**Table 1**). Regarding outcomes, babies from the Sz-HIE group took longer to become non-invasively ventilated and to reach full enteral feeds compared to those in the Sz-no HIE and no Sz-HIE groups (**Table S1a**). Neonates in the Sz-HIE had more severe levels of brain injury using the NICHD-NRN MRI scale than those without Sz (57% vs. 79%, p<0.001; **Table 1**)

**FIGURE 1.**
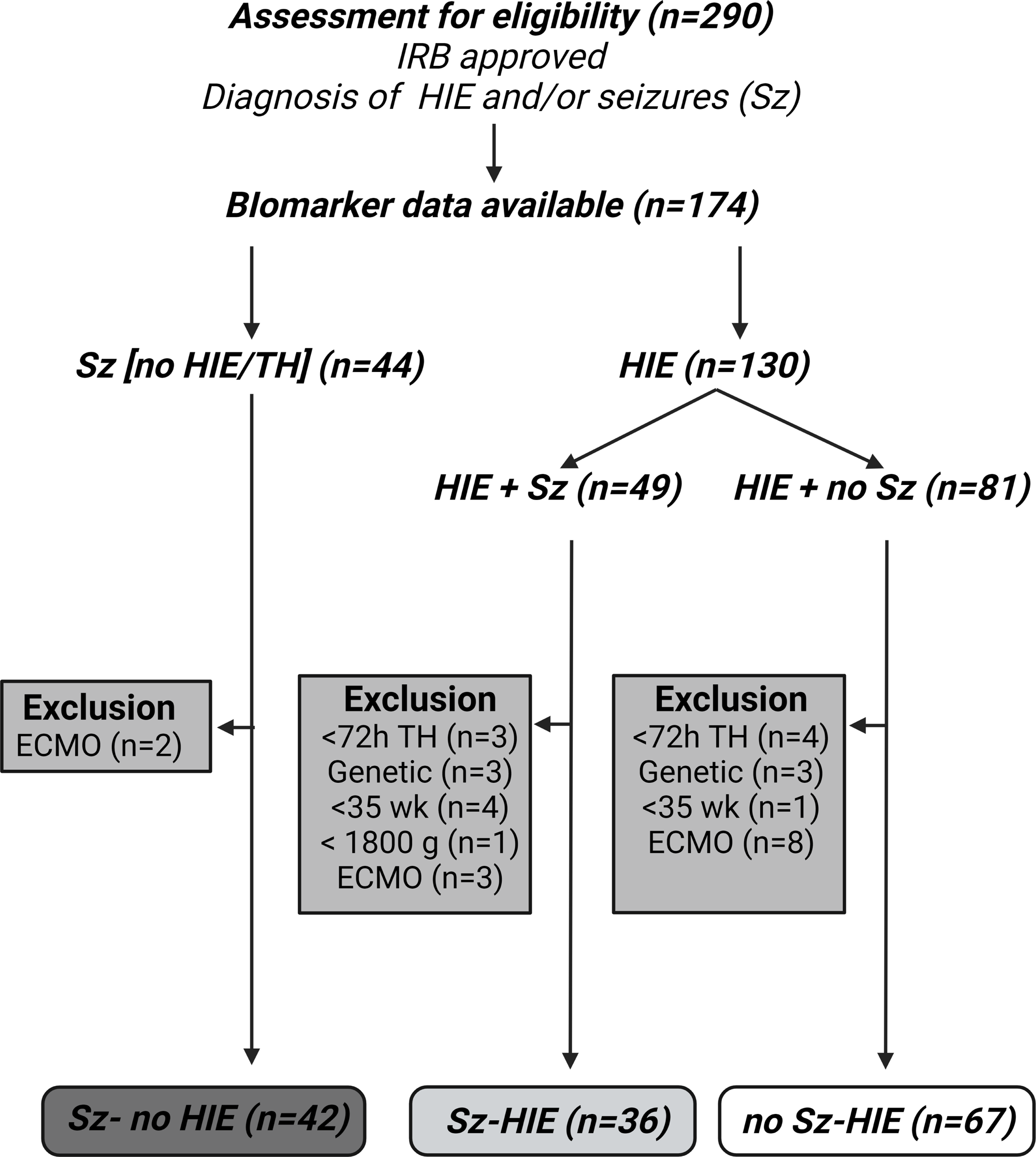
Patient allocation.

#### Serum biomarkers

At admission, GFAP levels were 2.1-fold higher in the Sz-HIE than in no Sz-HIE (p=0.007; **Fig 2A and Tables 2 & S2a**), while Tau and NF-L levels were similar (**Fig 2C & 2E**). Within the first 72h of life, Sz-no HIE and Sz-HIE groups showed a 3.7 and 2.3-fold higher GFAP (**Fig 2A**), 2.7 and 3.1-fold higher Tau (p<0.001; **Fig 2C**) and 1.9 and 2.9-fold higher NF-L (p=0.002; **Fig 2E**) levels than those in the no-Sz HIE group (**Tables 2 & S2a**) even after excluding neonates with stroke (**Tables 2 and S2b**) or those with mild encephalopathy (**Tables 2, S2c & S3**). Serum GFAP and Tau levels remained unchanged or even decreased during TH compared to admission in both HIE groups; however, serum NF-L levels continue to increase more robustly in the Sz-HIE group than the no Sz-HIE group (**Table S4**). Although after 72h of life, NF-L levels increased by 2.1-fold in no-Sz HIE neonates (**Tables 2 & S4**), NF-L levels in both Sz-no HIE and Sz-HIE groups were 3.8 and 2.8-fold higher than those in the no Sz-HIE group (**Table S2a**), respectively; even after excluding stroke cases (**Table S2b**) or mild encephalopathy (**Table S2c**). After the first 72h of life, Tau and NF-L levels remained higher in Sz-no HIE and Sz-HIE than in the no Sz-HIE group (**Fig 2; Tables 2 & S2a**) and these differences persist after removing stroke cases (**Tables 2 and S2b**). In HIE groups, Sarnat scores appear randomly distributed relative to the biomarker levels and any given time (**Fig S2**). Furthermore, excluding Sarnat scores <2 did not attenuate the differences in biomarkers identified between Sz-HIE and no Sz-HIE groups (**Tables 2, S2c & S3**).

**FIGURE 2.**
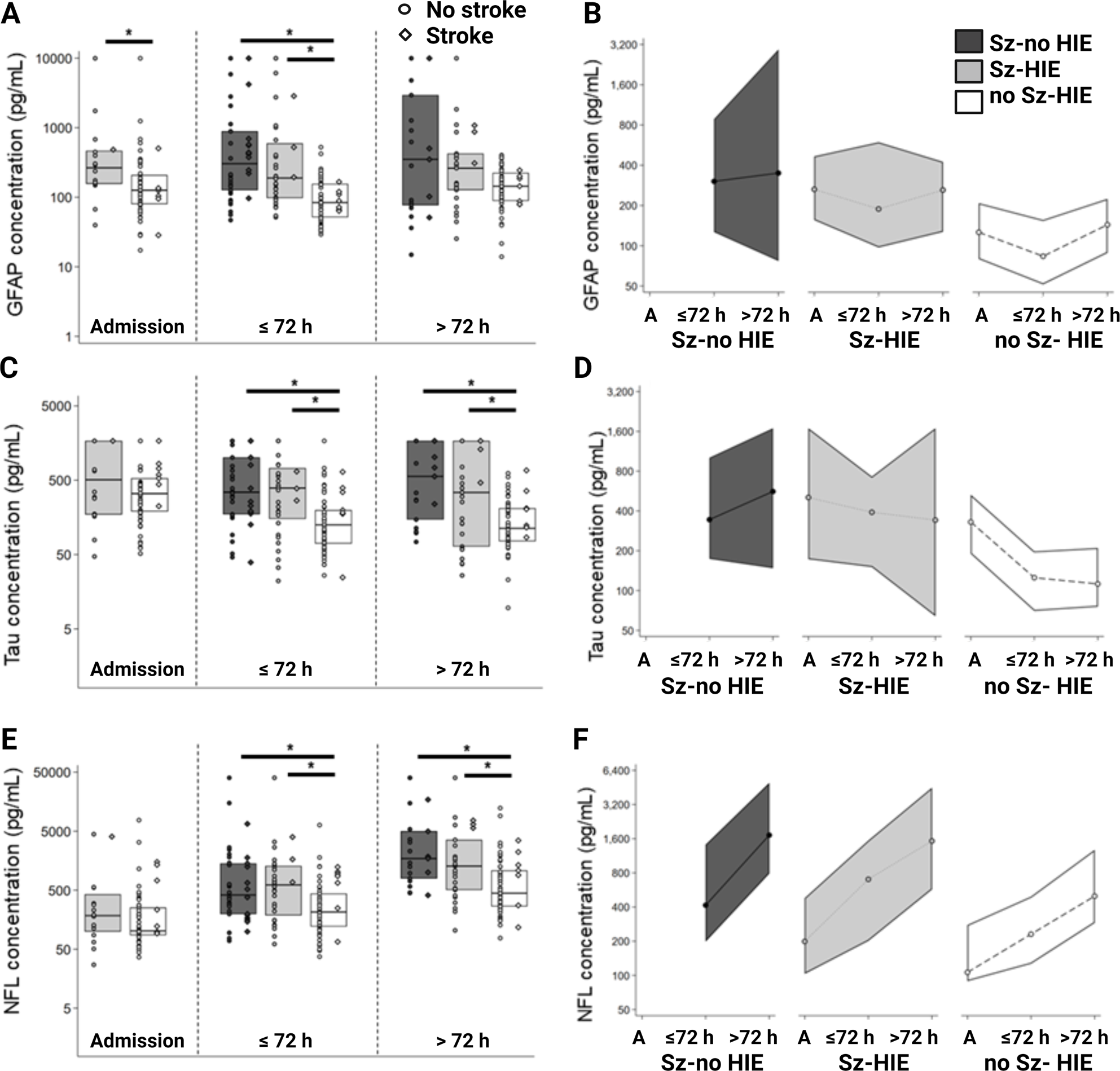
Box-plots of biomarker serum concentrations, by group (dark grey = Sz-no HIE; light grey = Sz-HIE; white = no Sz-HIE) and timepoint (Admission [A], ≤ 72h, and >72h [to 144h]) demonstrating neonates without (circle) and with (rhombus) diagnosis of stroke for (**A**) GFAP, (**C**) Tau and (**E**) NF-L. Boxes represent distribution from 25^th^ to 75^th^ percentile; solid line indicates the median. Longitudinal arrangements are presented in profile plot of unadjusted median biomarker concentrations (pg/mL) limited by IQR, by group and timepoint for (**B**) GFAP, (**D**) Tau, and (**F**) NF-L. **GFAP,** Glial fibrillary acidic protein; **h**, hours; **NF-L,** neurofilament light.

**Table 2.**
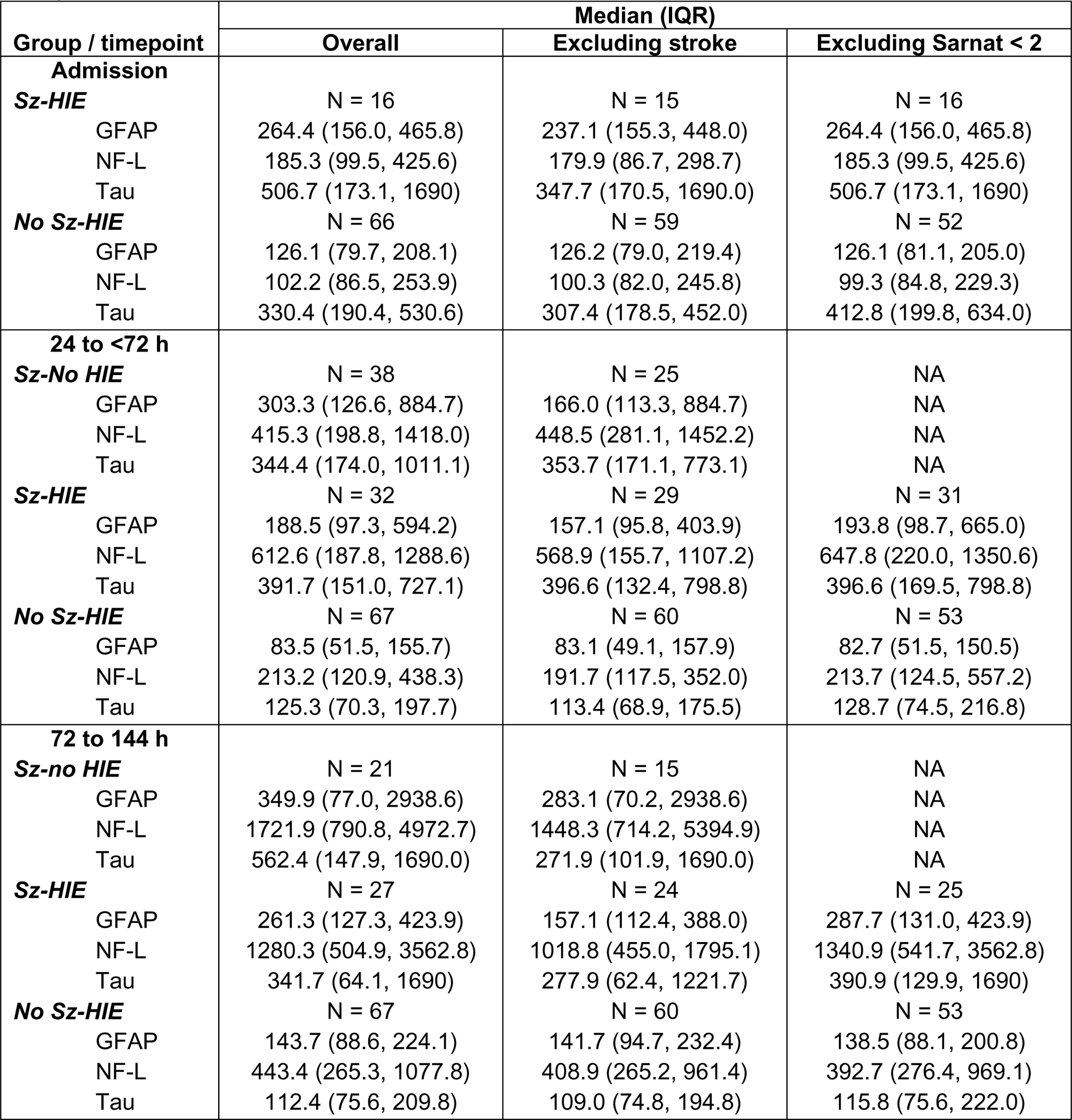
Descriptive statistics of serum GFAP, NF-L, and Tau concentrations (pg/mL), by group and timepoint.

| Group / timepoint | Median (IQR) |  |  |
| --- | --- | --- | --- |
|  | Overall | Excluding stroke | Excluding Sarnat < 2 |
| **Admission** |  |  |  |
| **Sz-HIE** | N = 16 | N = 15 | N = 16 |
| GFAP | 264.4 (156.0, 465.8) | 237.1 (155.3, 448.0) | 264.4 (156.0, 465.8) |
| NF-L | 185.3 (99.5, 425.6) | 179.9 (86.7, 298.7) | 185.3 (99.5, 425.6) |
| Tau | 506.7 (173.1, 1690) | 347.7 (170.5, 1690.0) | 506.7 (173.1, 1690) |
| **No Sz-HIE** | N = 66 | N = 59 | N = 52 |
| GFAP | 126.1 (79.7, 208.1) | 126.2 (79.0, 219.4) | 126.1 (81.1, 205.0) |
| NF-L | 102.2 (86.5, 253.9) | 100.3 (82.0, 245.8) | 99.3 (84.8, 229.3) |
| Tau | 330.4 (190.4, 530.6) | 307.4 (178.5, 452.0) | 412.8 (199.8, 634.0) |
| **24 to <72 h** |  |  |  |
| **Sz-No HIE** | N = 38 | N = 25 | NA |
| GFAP | 303.3 (126.6, 884.7) | 166.0 (113.3, 884.7) | NA |
| NF-L | 415.3 (198.8, 1418.0) | 448.5 (281.1, 1452.2) | NA |
| Tau | 344.4 (174.0, 1011.1) | 353.7 (171.1, 773.1) | NA |
| **Sz-HIE** | N = 32 | N = 29 | N = 31 |
| GFAP | 188.5 (97.3, 594.2) | 157.1 (95.8, 403.9) | 193.8 (98.7, 665.0) |
| NF-L | 612.6 (187.8, 1288.6) | 568.9 (155.7, 1107.2) | 647.8 (220.0, 1350.6) |
| Tau | 391.7 (151.0, 727.1) | 396.6 (132.4, 798.8) | 396.6 (169.5, 798.8) |
| **No Sz-HIE** | N = 67 | N = 60 | N = 53 |
| GFAP | 83.5 (51.5, 155.7) | 83.1 (49.1, 157.9) | 82.7 (51.5, 150.5) |
| NF-L | 213.2 (120.9, 438.3) | 191.7 (117.5, 352.0) | 213.7 (124.5, 557.2) |
| Tau | 125.3 (70.3, 197.7) | 113.4 (68.9, 175.5) | 128.7 (74.5, 216.8) |
| **72 to 144 h** |  |  |  |
| **Sz-no HIE** | N = 21 | N = 15 | NA |
| GFAP | 349.9 (77.0, 2938.6) | 283.1 (70.2, 2938.6) | NA |
| NF-L | 1721.9 (790.8, 4972.7) | 1448.3 (714.2, 5394.9) | NA |
| Tau | 562.4 (147.9, 1690.0) | 271.9 (101.9, 1690.0) | NA |
| **Sz-HIE** | N = 27 | N = 24 | N = 25 |
| GFAP | 261.3 (127.3, 423.9) | 157.1 (112.4, 388.0) | 287.7 (131.0, 423.9) |
| NF-L | 1280.3 (504.9, 3562.8) | 1018.8 (455.0, 1795.1) | 1340.9 (541.7, 3562.8) |
| Tau | 341.7 (64.1, 1690) | 277.9 (62.4, 1221.7) | 390.9 (129.9, 1690) |
| **No Sz-HIE** | N = 67 | N = 60 | N = 53 |
| GFAP | 143.7 (88.6, 224.1) | 141.7 (94.7, 232.4) | 138.5 (88.1, 200.8) |
| NF-L | 443.4 (265.3, 1077.8) | 408.9 (265.2, 961.4) | 392.7 (276.4, 969.1) |
| Tau | 112.4 (75.6, 209.8) | 109.0 (74.8, 194.8) | 115.8 (75.6, 222.0) |

#### Outcome data

After adjustment for BW, delivery mode and multiorgan failure, higher post-admission Tau (**Fig 3A**) or NF-L (**Fig 3B**) levels predicted longer time to reach full oral feeds in neonates from either Sz group (without or with HIE), but not in those from no Sz-HIE group. Serum levels for GFAP, Tau, and NF-L at admission also related directly to longer time to achieve full oral feeds in babies from the Sz-HIE, but not in those from the no-Sz-HIE group (**Fig S3**). Assess of the magnitude of importance of clinical variables and biomarkers as features relative to neonatal outcomes was performed using SHAP analysis. The SHAP modeling was good to excellent in predicting impaired oral feedings (**Fig 3C & 3D**). Multi-organ dysfunction, BW, and NRFHT were among the most important clinical features influencing time to full feeds. The direct relations of Tau, NF-L, and GFAP were observed in all models for the Sz groups (**Fig 3C & 3D**). Ordinal logistic regression models adjusted for covariates (including Sz) showed that doubling in NF-L and Tau by 72 to 144h of life was associated with a 2-fold increase in the odds to have a worse brain injury level in the NICHD-NRN MRI scale. Similarly, adjusted logistic regression models of significant brain injury (score ≥1B), higher Tau levels at 72 to 144h were associated with higher odds of significant brain injury (**Table 3**). When evaluating Sz as a clinical feature in babies with HIE (combining Sz-HIE and no Sz-HIE, **Fig 3E**), Sz was the most important feature in SHAP analysis modeling significant brain injury (score ≥1B in NICHD-NRN MRI scale), with multiorgan failure being the second. Higher Tau and NF-L were also associated with worse brain injury in MRI. Similarly, SHAP analysis including only the Sz-HIE group also showed direct association between higher NF-L and additionally GFAP, with significant brain injury in this group.

**FIGURE 3.**
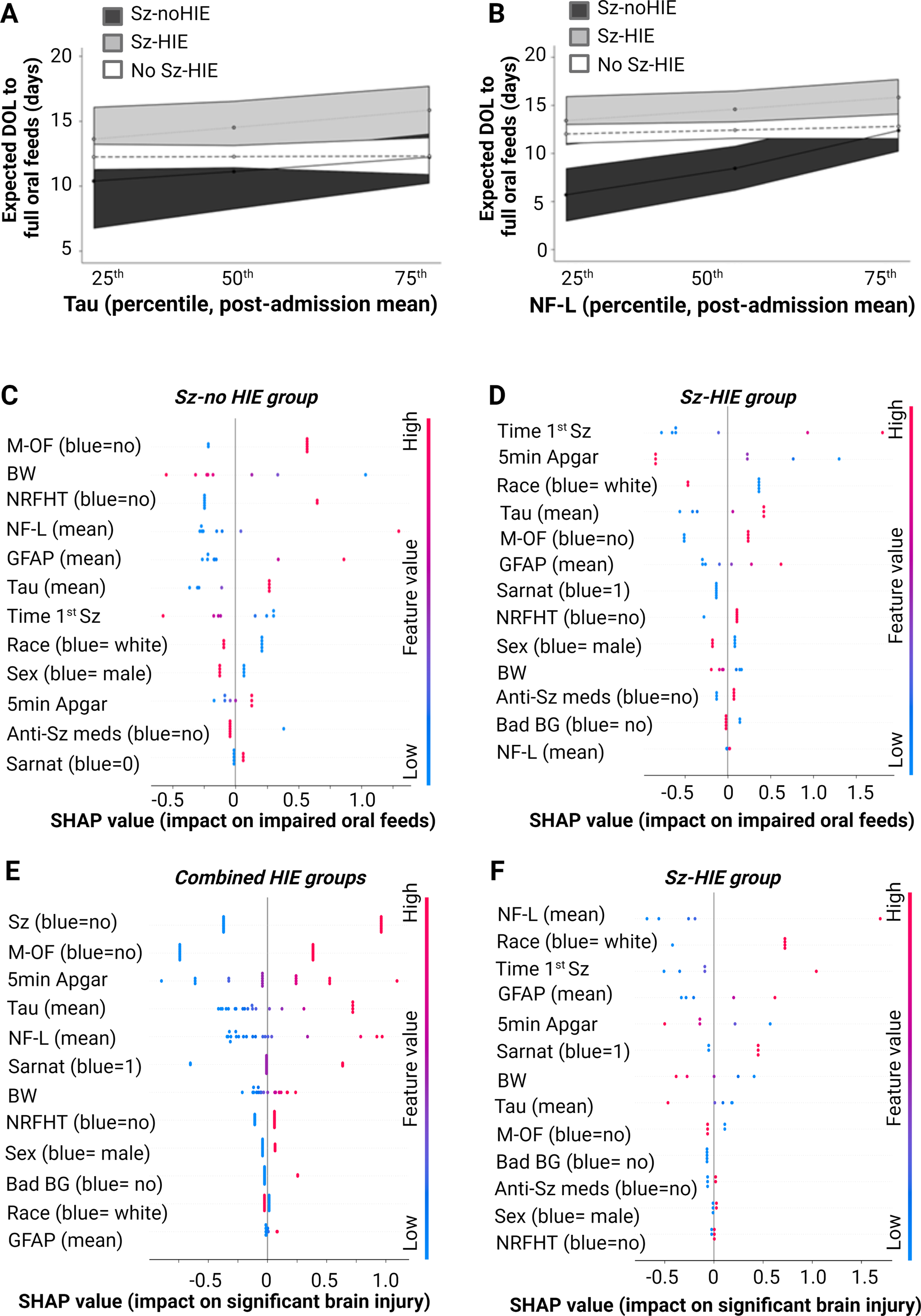
Adjusted expected days of life (DOL) to full oral feed, by group and percentile of biomarker concentration (pg/mL) post-natally for (**A**) Tau and (**B**) NF-L. Estimates are adjusted for birthweight, delivery mode and multi-organ failure, and biomarker concentration. Shapley Additive explanations (SHAP) value plots provide a hierarchical organization of various features included in the model to predict the likelihood of impaired oral feedings in the (**C**) Sz and (**D**) Sz-HIE groups. Similarly, SHAP value plots organized features included in the model to predict significant brain injury using NICHD-NRN MRI scale system greater than 1A level as “significant” for the whole HIE cohort (**E**) and the Sz-HIE group (**F**). Red and blue dots represent higher and lower values of the feature and positive (right from zero) and negative (left from zero) SHAP value represent the degree by which increased or decreases the risk of the outcome. **NF-L,** neurofilament light; **NRFHT,** non-reassuring fetal heart tracing, **Sz**, seizure.

**Table 3.** Adjusted odds ratios (95% confidence intervals) for significant MRI-based brain injury (NICHD NRN score > 1A) per doubling in brain injury biomarker, among those with HIE.

| **Biomarker and timepoint** | **Cumulative OR (95% CI)**<br>**All brain injury<sup>a</sup>** | **OR (95% CI)**<br>**Significant brain injury<sup>b</sup>** |
| --- | --- | --- |
| **GFAP** |  |  |
| Admission, n = 68 | 0.94 (0.63, 1.41)<br>P = 0.77 | 0.82 (0.41, 1.66)<br>P = 0.58 |
| 24 to <72 hours, n = 78<sup>c</sup> | 1.69 (0.93, 3.05)<br>P = 0.08 | 1.26 (0.53, 3.02)<br>P = 0.60 |
| 72 to 144 hours, n = 78<sup>c</sup> | 1.47 (0.71, 3.05)<br>P = 0.30 | 1.75 (0.62, 4.91)<br>P = 0.29 |
| **NF-L** |  |  |
| Admission, n = 68 | 1.24 (0.85, 1.80)<br>P = 0.26 | 0.99 (0.58, 1.69)<br>P = 0.99 |
| 24 to <72 hours, n = 78<sup>c</sup> | 0.89 (0.48, 1.63)<br>P = 0.70 | 0.62 (0.27, 1.42)<br>P = 0.26 |
| 72 to 144 hours, n = 78<sup>c</sup> | **2.00 (1.01, 3.98)**<br>**P = 0.046** | 2.14 (0.90, 5.07)<br>P = 0.08 |
| **Tau** |  |  |
| Admission, n = 68 | **1.57 (1.02, 2.40)**<br>**P = 0.040** | 1.07 (0.63, 1.81)<br>P = 0.82 |
| 24 to <72 hours, n = 78<sup>c</sup> | 0.98 (0.50, 1.92)<br>P = 0.90 | 0.43 (0.16, 1.10)<br>P = 0.08 |
| 72 to 144 hours, n = 78<sup>c</sup> | **2.12 (1.02, 4.40)**<br>**P = 0.044** | **2.70 (1.03, 7.09)**<br>**P = 0.044** |

Note: All models were adjusted for birthweight, delivery mode (vaginal vs. cesarean-section/other), multi-organ failure, and seizures.
<sup>a</sup> Estimated using ordinal logistic regression; estimates are cumulative odds ratios for being in a higher brain injury severity category per doubling in brain injury biomarker.
<sup>b</sup> Estimated using penalized logistic regression; estimates are odds ratios for being in the significant brain injury category per doubling in brain injury biomarker.
<sup>c</sup> Models additionally co-adjusted for biomarker concentrations at 24 to <72 hours postnatally and 72 to 144 hours postnatally.

## DISCUSSION

Seizures are a marker of severity of HI brain injury and thus, are used as an indication of at least moderate HIE. However, one of the most urgent yet debated questions has been whether Sz may worsen brain injury in HIE and other cerebral pathologies. Here, we present interpretations from CNS-specific serum biomarkers, which after adjustment for confounders, support the hypothesis that astrocytic activation (GFAP), neuronal injury and death (Tau), and axonal degeneration (NF-L) occur more robustly in the setting of Sz, even without HIE. Longer time to reach full oral feeds, a well-studied surrogate of neurological recovery,^17^ and significant brain injury in MRI by NICHD-NRN scale^18^, which relates to worst neurodevelopmental impairment, are both associated with higher levels of these biomarkers. Based on our analysis, the temporal utility separates GFAP and Tau as earlier biomarkers, while NF-L as a delayed biomarker.

GFAP, Tau and NF-L are nervous system-specific biomarkers. While Tau and GFAP reach circulation upon injury (and probably death) of neurons and astrocytes, respectively,^15,19^ NF-L is an axonal cytoskeletal protein, which reaches circulation upon axonal degeneration. Thus, the temporal relationship between these biological processes supports the contention that GFAP and Tau would peak earlier in serum than NF-L, as axonal degeneration follows initial astrocytic activation (GFAP) and neuronal injury (Tau). Supporting this hypothesis, we document that in HIE neonates without Sz, NF-L increased over time, while GFAP and Tau declined over time. In contrast, supporting ongoing brain injury (and cell death) following HI insult in neonates who developed Sz, we find persistently increased GFAP and Tau levels along with rise in NF-L levels. These differences persist even after neonates with strokes are removed, making a primary biological effect from Sz plausible.

Despite its observer-bias nature,^12^ the Sarnat score remains among the most brain-specific indicators available to assess clinical NE.^1^ We and others have confirmed the association between Sarnat scores and circulating Tau and GFAP.^15,20–23^ We have recently shown, after adjustments and stratification by Sarnat score, that neonates suffering of HIE with Sarnat score of 3 have higher GFAP within the first 24h of life, while Tau levels begin to increase during the next 72h (TH period).^20^ In adjusted models, the associations between Sarnat scores with Tau and GFAP become stronger after TH,^20^ may imply persistence or even worsen brain injury after rewarming in more severely encephalopathic neonates as shown in several RCTs.^11,24^ Using the ultrasensitive MSD^®^ platform approach, we now measured more reliably lower levels for GFAP, Tau, and additionally NF-L. Here, we identified that neither GFAP, Tau nor NF-L serum levels changed by Sarnat score after adjustments in HIE infants with no Sz. Conversely, all biomarkers increased in those HIE neonates with Sz in an incremental manner relative to higher Sarnat scores. Thus, Sz may worsen brain injury, particularly in neonates with higher Sarnat scores within the first 6h of life.

The use of phenobarbital, as the known first line anti-seizure medication in neonates, represented over 90% of all treatments used in both Sz groups. Overall, the use of anti-seizure medications appears to have a weak influence in the regression models to predict significant injury in brain MRI or short-term outcomes (delayed oral feeding); however, the stability of the modeling is decreased due to the low frequency of no treatment in those infants demonstrating confirmed Sz. In this cohort from 2009 to 2019, 79.2% and 63.6% of neonates from the Sz-no HIE and Sz HIE groups, respectively, were discharged home with anti-seizure medications, representing the management trends during that study period. Thus, associations between biomarkers and medications at discharge did not represent a biological process, reason why this analysis was not pursued. Nevertheless, the use of anti-seizure medications in neonates, specifically phenobarbital, as a modulator of these associations needs further study considering its known pro-apoptotic effects.^25,26^

Although, SHAP analysis confirms the high impact of all 3 biomarkers in the prediction of impaired transition to full oral feeds and significant brain injury in neonates with Sz, along with clinical features such as higher severity of illness (presence of multiorgan failure), NRFHT, lower 5 min Apgar score, male sex and non-white race, we must acknowledge the limitations of our study. The influence of the brain-blood-barrier (BBB) microstructural disruption and recovery during and after TH in the detection of circulating biomarkers remains unexplored.^27^ Thus, the lack of decline in GFAP and Tau in HIE neonates with Sarnat scores ≥ 2 may be the result of persistent BBB disruption, although this mechanism would not explain the robust rise in NF-L, suggesting instead ongoing axonal degeneration. Seizure burden, reflecting the severity and the degree of ictal activity refractory to treatment, would have allowed a dose-response analysis, providing more support for direct causation. However, this degree of granularity of the EEG data was not available in this cohort. Although previous studies have not identified associations between timing of TH initiation and cerebral autoregulation,^28^ injury on brain MRI, or neurodevelopmental outcomes,^29^ time to TH initiation may still influence the biomarker trajectories. Additionally clinical practice drift occurring during this study is possible but difficult to isolate. Although no difference was found between neonates with HIE with and without biomarker data (**Table S1b**), selection bias is still possible, since a small subset of very sick neonates lacked samples for analysis and their parents were not approached for consent. However, if we assume a monotonic relation between brain injury biomarkers and degree or severity of illness, then the exclusion of the most critically ill neonates would bias our observed associations towards the null, and therefore our findings are likely conservative.

## Conclusions

Relative to neonates without Sz, concentrations of CNS-specific biomarkers linked to brain injury are higher in those developing Sz. Concentrations were similar between neonates in the Sz and Sz-HIE groups providing support to the notion that Sz may worsen brain injury in neonates with HIE and are associated with worse outcomes, particularly in those with higher Sarnat scores.

## ACKNOWLEDGEMENTS

We thank the families of the extremely sick neonates included in this study for their willingness to participate. We also thank the nursing and ancillary staff at Johns Hopkins Hospital NICU and core laboratory for their support in the collection of the samples. Lastly, we thank Ms. Jie Zhu, research specialist at ADE lab for the processing of samples.

## FINANCIAL STATEMENT

Supported by National Institutes of Health RO1 HD110091 (KBA, JK, CP, GJG, AT, EMG, ADE, FJN, RC-V); RO1 NS126549 (GJG, FJN, RC-V); U01NS114144 (CD, MS, GS, JW) and the Thomas Wilson Foundation (RC-V).

## AUTHOR CONTRIBUTIONS

Conceptualization and design of the study (KBA, KJ, ADE, FJN, RC-V); methodology (KBA, KJ, CP, AT, EMG, CES, ADE, FJN, RC-V); supervision and oversight (ADE, FJN, RC-V); funding acquisition (KBA, FJN, ADE, RC-V, CC); data acquisition (all authors), formal data analysis (KBA, JK, RCV, HK); resources (KBA, KJ, CP, GJG, AT, EMG, CES, CD, CC, MS, GS, JW, ADE, FJN, RC-V); drafting of significant portions of manuscript, tables and figures (KBA, JK, RC-V, HK); and review and editing of final manuscript (all authors).

## DATA AVAILABILITY

The information summarized in this article contains deidentified protected health information (PHI). Consequently, the data is not available to be shared due to privacy and confidentiality considerations.

Additional requests for information regarding the data should be directed to the corresponding author.

## SUPPLEMENTAL FIGURE LEGENDS

**FIGURE S1.** Box-plots of biomarker serum concentrations, by group (dark grey = Sz-no HIE; light grey = Sz-HIE; white = no Sz-HIE) and timepoint (Admission [A], ≤ 72h, and >72h [to 144h]) demonstrating neonates for (**A1**) GFAP, (**B1**) Tau and (**C1**) NF-L. Boxes represent distribution from 25^th^ to 75^th^ percentile; solid line indicates median. Longitudinal arrangements are presented in profile plot of unadjusted median (IQR) brain injury biomarker concentrations (pg/mL) limited by IQR, by group and timepoint for (**A2**) GFAP, (**B2**) Tau, and (**C2**) NF-L. **GFAP,** Glial fibrillary acidic protein; **hrs**, hours; **NF-L,** neurofilament light.

**FIGURE S2.** Box-plots of biomarker serum concentrations, by group and timepoint demonstrating neonates by Sarnat score. (**A**) GFAP, (**B**) NF-L, and (**C**) Tau. Boxes represent distribution from 25^th^ to 75^th^ percentile; solid line indicates median. **GFAP,** Glial fibrillary acidic protein; **hrs**, hours; **NF-L,** neurofilament light.

**FIGURE S3.** Adjusted expected days of life (DOL) to full oral feed, by group and percentile of biomarker concentration (pg/mL) at admission for (**A**) GFAP, (**B**) Tau and (**C**) NF-L. Estimates are adjusted for birthweight and delivery mode (vaginal vs. cesarean-section/other), multi-organ failure. **GFAP,** Glial fibrillary acidic protein; **h**, hours; **NF-L,** neurofilament light.

## REFERENCES

1. Douglas-Escobar M, Weiss MD. Hypoxic-ischemic encephalopathy: a review for the clinician. JAMA Pediatr. 2015;169:397–403. doi: 10.1001/jamapediatrics.2014.3269

2. Jantzie LL, Getsy PM, Denson JL, Firl DJ, Maxwell JR, Rogers DA, Wilson CG, Robinson S. Prenatal Hypoxia-Ischemia Induces Abnormalities in CA3 Microstructure, Potassium Chloride Co-Transporter 2 Expression and Inhibitory Tone. Front Cell Neurosci. 2015;9:347. doi: 10.3389/fncel.2015.00347

3. Khazipov R, Khalilov I, Tyzio R, Morozova E, Ben-Ari Y, Holmes GL. Developmental changes in GABAergic actions and seizure susceptibility in the rat hippocampus. Eur J Neurosci. 2004;19:590–600. doi: 10.1111/j.0953-816x.2003.03152.x

4. Pond BB, Berglund K, Kuner T, Feng G, Augustine GJ, Schwartz-Bloom RD. The chloride transporter Na(+)-K(+)-Cl-cotransporter isoform-1 contributes to intracellular chloride increases after in vitro ischemia. J Neurosci. 2006;26:1396–1406. doi: 10.1523/JNEUROSCI.1421-05.2006

5. Jensen FE. Neonatal seizures: an update on mechanisms and management. Clin Perinatol. 2009;36:881–900, vii. doi: 10.1016/j.clp.2009.08.001

6. Shao LR, Habela CW, Stafstrom CE. Pediatric Epilepsy Mechanisms: Expanding the Paradigm of Excitation/Inhibition Imbalance. Children (Basel). 2019;6. doi: 10.3390/children6020023

7. Basti C, Maranella E, Cimini N, Catalucci A, Ciccarelli S, Del Torto M, Di Luca L, Di Natale C, Mareri A, Nardi V, et al. Seizure burden and neurodevelopmental outcome in newborns with hypoxic-ischemic encephalopathy treated with therapeutic hypothermia: A single center observational study. Seizure. 2020;83:154–159. doi: 10.1016/j.seizure.2020.10.021

8. Lin YK, Hwang-Bo S, Seo YM, Youn YA. Clinical seizures and unfavorable brain MRI patterns in neonates with hypoxic ischemic encephalopathy. Medicine (Baltimore). 2021;100:e25118. doi: 10.1097/MD.0000000000025118

9. Kharoshankaya L, Stevenson NJ, Livingstone V, Murray DM, Murphy BP, Ahearne CE, Boylan GB. Seizure burden and neurodevelopmental outcome in neonates with hypoxic-ischemic encephalopathy. Dev Med Child Neurol. 2016;58:1242–1248. doi: 10.1111/dmcn.13215

10. Davidson JO, Bennet L, Gunn AJ. Evaluating anti-epileptic drugs in the era of therapeutic hypothermia. Pediatr Res. 2019;85:931–933. doi: 10.1038/s41390-019-0319-6

11. Shankaran S, Laptook AR, Ehrenkranz RA, Tyson JE, McDonald SA, Donovan EF, Fanaroff AA, Poole WK, Wright LL, Higgins RD, et al. Whole-body hypothermia for neonates with hypoxic-ischemic encephalopathy. N Engl J Med. 2005;353:1574–1584. doi: 10.1056/NEJMcps050929

12. Sarnat HB, Sarnat MS. Neonatal encephalopathy following fetal distress. A clinical and electroencephalographic study. Arch Neurol. 1976;33:696–705. doi: 10.1001/archneur.1976.00500100030012

13. Murray DM, Bala P, O’Connor CM, Ryan CA, Connolly S, Boylan GB. The predictive value of early neurological examination in neonatal hypoxic-ischaemic encephalopathy and neurodevelopmental outcome at 24 months. Dev Med Child Neurol. 2010;52:e55–59. doi: 10.1111/j.1469-8749.2009.03550.x

14. Shankaran S, Pappas A, McDonald SA, Laptook AR, Bara R, Ehrenkranz RA, Tyson JE, Goldberg R, Donovan EF, Fanaroff AA, et al. Predictive value of an early amplitude integrated electroencephalogram and neurologic examination. Pediatrics. 2011;128:e112–120. doi: 10.1542/peds.2010-2036

15. Dietrick B, Molloy E, Massaro AN, Strickland T, Zhu J, Slevin M, Donoghue V, Sweetman D, Kelly L, O’Dea M, et al. Plasma and Cerebrospinal Fluid Candidate Biomarkers of Neonatal Encephalopathy Severity and Neurodevelopmental Outcomes. J Pediatr. 2020;226:71–79 e75. doi: 10.1016/j.jpeds.2020.06.078

16. Hornung RW. Estimation of Average Concentration in the Presence of Nondetectable Values. Applied Occupational and Environmental Hygiene. 1990;5:46–51. doi: 10.1080/1047322X.1990.10389587

17. Graham EM, Ruis KA, Hartman AL, Northington FJ, Fox HE. A systematic review of the role of intrapartum hypoxia-ischemia in the causation of neonatal encephalopathy. Am J Obstet Gynecol. 2008;199:587–595. doi: 10.1016/j.ajog.2008.06.094

18. Shankaran S, Barnes PD, Hintz SR, Laptook AR, Zaterka-Baxter KM, McDonald SA, Ehrenkranz RA, Walsh MC, Tyson JE, Donovan EF, et al. Brain injury following trial of hypothermia for neonatal hypoxic-ischaemic encephalopathy. Arch Dis Child Fetal Neonatal Ed. 2012;97:F398–404. doi: 10.1136/archdischild-2011-301524

19. Graham EM, Burd I, Everett AD, Northington FJ. Blood Biomarkers for Evaluation of Perinatal Encephalopathy. Front Pharmacol. 2016;7:196. doi: 10.3389/fphar.2016.00196

20. Chavez-Valdez R, Miller S, Spahic H, Vaidya D, Parkinson C, Dietrick B, Brooks S, Gerner GJ, Tekes A, Graham EM, et al. Therapeutic Hypothermia Modulates the Relationships Between Indicators of Severity of Neonatal Hypoxic Ischemic Encephalopathy and Serum Biomarkers. Front Neurol. 2021;12:748150. doi: 10.3389/fneur.2021.748150

21. Lv HY, Wu SJ, Gu XL, Wang QL, Ren PS, Ma Y, Peng LY, Jin LH, Li LX. Predictive Value of Neurodevelopmental Outcome and Serum Tau Protein Level in Neonates with Hypoxic Ischemic Encephalopathy. Clin Lab. 2017;63:1153–1162. doi: 10.7754/Clin.Lab.2017.170103

22. Massaro AN, Wu YW, Bammler TK, Comstock B, Mathur A, McKinstry RC, Chang T, Mayock DE, Mulkey SB, Van Meurs K, et al. Plasma Biomarkers of Brain Injury in Neonatal Hypoxic-Ischemic Encephalopathy. J Pediatr. 2018;194:67–75 e61. doi: 10.1016/j.jpeds.2017.10.060

23. Takahashi K, Hasegawa S, Maeba S, Fukunaga S, Motoyama M, Hamano H, Ichiyama T. Serum tau protein level serves as a predictive factor for neurological prognosis in neonatal asphyxia. Brain Dev. 2014;36:670–675. doi: 10.1016/j.braindev.2013.10.007

24. Gluckman PD, Wyatt JS, Azzopardi D, Ballard R, Edwards AD, Ferriero DM, Polin RA, Robertson CM, Thoresen M, Whitelaw A, et al. Selective head cooling with mild systemic hypothermia after neonatal encephalopathy: multicentre randomised trial. Lancet. 2005;365:663–670. doi: 10.1016/S0140-6736(05)17946-X

25. Bittigau P, Sifringer M, Genz K, Reith E, Pospischil D, Govindarajalu S, Dzietko M, Pesditschek S, Mai I, Dikranian K, et al. Antiepileptic drugs and apoptotic neurodegeneration in the developing brain. Proc Natl Acad Sci U S A. 2002;99:15089–15094. doi: 10.1073/pnas.222550499

26. Forcelli PA, Kim J, Kondratyev A, Gale K. Pattern of antiepileptic drug-induced cell death in limbic regions of the neonatal rat brain. Epilepsia. 2011;52:e207–211. doi: 10.1111/j.1528-1167.2011.03297.x

27. Saunders NR, Dziegielewska KM, Mollgard K, Habgood MD. Recent Developments in Understanding Barrier Mechanisms in the Developing Brain: Drugs and Drug Transporters in Pregnancy, Susceptibility or Protection in the Fetal Brain? Annu Rev Pharmacol Toxicol. 2019;59:487–505. doi: 10.1146/annurev-pharmtox-010818-021430

28. Gilmore MM, Tekes A, Perin J, Parkinson C, Spahic H, Chavez-Valdez R, Northington FJ, Lee JK. Later cooling within 6 h and temperatures outside 33-34 degrees C are not associated with dysfunctional autoregulation during hypothermia for neonatal encephalopathy. Pediatr Res. 2021;89:223–230. doi: 10.1038/s41390-020-0876-8

29. Guillot M, Philippe M, Miller E, Davila J, Barrowman NJ, Harrison MA, Ben Fadel N, Redpath S, Lemyre B. Influence of timing of initiation of therapeutic hypothermia on brain MRI and neurodevelopment at 18 months in infants with HIE: a retrospective cohort study. BMJ Paediatr Open. 2019;3:e000442. doi: 10.1136/bmjpo-2019-000442

